# Pre-diagnostic loss to follow-up in an active case-finding TB program: a mixed-methods study from rural Bihar, India

**DOI:** 10.1101/19005199

**Authors:** Tushar Garg, Vivek Gupta, Dyuti Sen, Madhur Verma, Miranda Brouwer, Rajeshwar Mishra, Manish Bhardwaj

**Author notes:** ADDRESS FOR CORRESPONDENCE Tushar Garg, Innovators In Health, #176, Ground Floor, Road #2, New Pataliputra Colony, Patna – 800013. Bihar. India.

## Abstract

**background:** Despite active case-finding (ACF) identifying more presumptive and confirmed TB cases, high pre-diagnostic loss to follow-up (PDLFU) among presumptive TB cases referred for diagnostic test remains a concern. We aimed to quantify the PDLFU, and identify the barriers and enablers in undergoing a diagnostic evaluation in an ACF program implemented in 1.02 million rural population in the Samastipur district of Bihar, India.

**methods:** During their routine work, Accredited Social Health Activists (ASHA, a community health worker or CHW), informal providers, and community laypersons identified people at risk of TB, and referred them to the program. A field coordinator (FC) screened them for TB symptoms at the patient’s home. The identified presumptive TB cases were accompanied by the CHW to a designated government facility for diagnostics. Those with a confirmed TB diagnosis were put on treatment by the CHW and followed-up till treatment completion. All services were provided free of cost and patients were supported throughout the care pathway, including a transport allowance. We analyzed programmatically collected data, conducted in-depth interviews with patients, and focus group discussions with the CHWs and FCs in an explanatory mixed-methods design.

**results:** A total of 11146 presumptive TB cases were identified from January 2018 to December 2018, out of which 4912 (44.1%) underwent a diagnostic evaluation. The key enablers were CHW accompaniment and support in addition to the free TB services in the public sector. The major barriers identified were transport challenges, deficient family and health provider support, and poor services in the public system.

**conclusion:** If we are to find missing cases, the health system needs urgent reform, and diagnostic services need to be patient-centric. A strong patient support system engaging all stakeholders and involvement of CHWs in routine TB care is an effective solution.

**STRENGTHS AND LIMITATIONS OF THIS STUDY:**

1. First such study to explore the reasons for pre-diagnostic loss to follow-up
2. A mixed-method design including the views of both patients and community health workers
3. Uses operational data from a routine programmatic setting at an NGO site
4. No record of the actual number of people screened intuitively before being referred to the program.
5. No record of patients accessing diagnostics in private sector and those completing the diagnostic process.

## INTRODUCTION

The End TB strategy targets an 80% reduction in TB incidence by 2030.[1] However, as many as 4.1 million (39%) patients globally are not notified, indicating a mix of under-reporting and under-diagnosis.[2] Of these, India alone has about a million.[3,4] Analysis of the cascade of care can effectively quantify the gaps in successive steps of TB care pathway and informs the TB programs of potential interventions.[4] The Indian public sector cascade and the South African cascade estimate a loss to follow up (LFU) at test access of 28% and 5% respectively out of incident cases.[5,6] Similar approaches can be used to evaluate case-finding programs involving systematic TB screening.[5]

Historically, most TB patients were identified through passive case-finding in which patients self-referred to a provider for a diagnostic evaluation.[7] It may lead to delays in diagnosis and continued transmission in the community.[8] The known major challenges in diagnosis are poor geographical and financial access to healthcare, fewer symptoms delay seeking healthcare, and failure to test when people do present at the facility.[9]

Diverse active case finding (ACF) strategies have demonstrated increased TB case detection with early diagnosis.[7,10,11] Nonetheless, multiple ACF interventions have reported high PDLFU. A study from India reported only 22% of the people with presumptive TB reaching a microscopy center, while an ACF program in Myanmar reported only 51.4% of patients with abnormal chest X-ray (CXR) getting their sputum examined.[12,13] A recent study of a TB program in a district of Zimbabwe reported a PDLFU of 25% with patient’s residence, HIV status and type of health facility emerging as significant risk factors.[14] Although these studies rely on routinely collected programmatic data, there has been no study exploring the reasons for the high PDLFU and the perspectives of associated stakeholders.

We aimed to identify the PDLFU, and assess the enablers and barriers for diagnostic evaluation in a community-based ACF program during 2018 in Bihar, India. The specific objectives were: 1) to assess proportion of people screened among community referrals, 2) to assess number of individuals with presumptive TB and their characteristics, 3) to assess proportion and characteristics of PDLFU patients, and identify risk factors, and 4) to understand the reasons for the PDLFU, including barriers and enablers to access the diagnostic evaluation.

## METHODS

### study design

We used an explanatory mixed-methods study design, where the quantitative phase (cohort analysis) was followed by a qualitative phase (descriptive design).[15]

### study setting

#### General setting

The study was conducted in the Sarairanjan, Bibhutipur, and Ujiarpur blocks (equivalent to TB Unit or TU) of Samastipur district in Bihar with a combined population of 1,021,483. Located in the Indo-Gangetic plain, agriculture was the main occupation of the population. The highest earning member in 69.8% of the households earned less than INR 60,000 annually (∼USD 852) and 63.5% of the population was reported literate.[16] Females had 20% lesser literacy rate compared to males, and infant mortality rate (IMR) was 60 for females against that of 48 for males.[17] The public TB case notification rate in the district was 55 per 100,000 population (2017) and the pre-treatment LFU was 25% (2017).[18]

The population was serviced by 12 primary health centres (PHC) under the Public Health System (PHS).[19] Within the Revised National TB Control Program (RNTCP), 4 designated microscopy centres (DMC) run by laboratory technicians provided sputum microscopy. A Senior Treatment Supervisor (STS) at the TU level managed the program and a Senior Treatment Laboratory Supervisor (STLS) oversaw a group of DMCs.

The last-mile TB care services were provided by a Community Health Worker (called ASHA or Accredited Social Health Activist). Initiated under the National Health Mission (NHM) in 2005, an ASHA is a literate woman of the village and assigned for every 1000 population. Her key responsibility is to be a facilitator, mobilizer, and community service provider for various public health programs.[20] Under the RNTCP, ASHA received activity-based remuneration: INR 1000 (∼ USD 14) for DS-TB treatment support, and INR 5000 (∼ USD 72) for DR-TB treatment support.[21]

Anganwadi workers (AWW) are community health workers under the Integrated Child Development Services Scheme and receive a fixed monthly honorarium. Through Angadwadi Centres, they provide early childhood care and education for children up to 6 years.[22]

#### Specific setting

We implemented a TB REACH Wave 5-funded novel, community-based ACF project in collaboration with the RNTCP in the above population. The key aspects were: 1) community referral of people at risk of TB, 2) symptom-based screening at patient’s doorstep, 3) ASHA-accompanied diagnostic evaluation, 4) training to ASHA, 5) extensive stakeholder engagement, and 6) engaging multiple community members, including AWW and informal providers (also known as RMP or Registered Medical Practitioner).

After an initial dialogue, ASHA, AWW, RMPs and the community were asked to refer people who may have TB. They relied on their existing knowledge of the community and looked for people at risk of TB during their routine work and social activities. These community referrals were reported to a field coordinator (FC) of the project, who screened them at their home for TB symptoms systematically. All presumptive TB cases were referred for a sputum microscopy and chest x-ray (CXR). All people with a positive smear, or people with a negative smear but abnormal CXR, or people with a negative smear but clinically suspect were offered a GeneXpert test using sputum collection and transport. ASHAs assisted presumptive TB patients in reaching the diagnostic centres and accompanied them through the process. Patients received a transport allowance and diagnostic tests were free of cost. If the PHS physician confirmed TB diagnosis, treatment was initiated under ASHA’s supervision. The ASHA delivered drugs to the patient, and supported them till treatment completion.[Figure 1]

**Figure 1:**
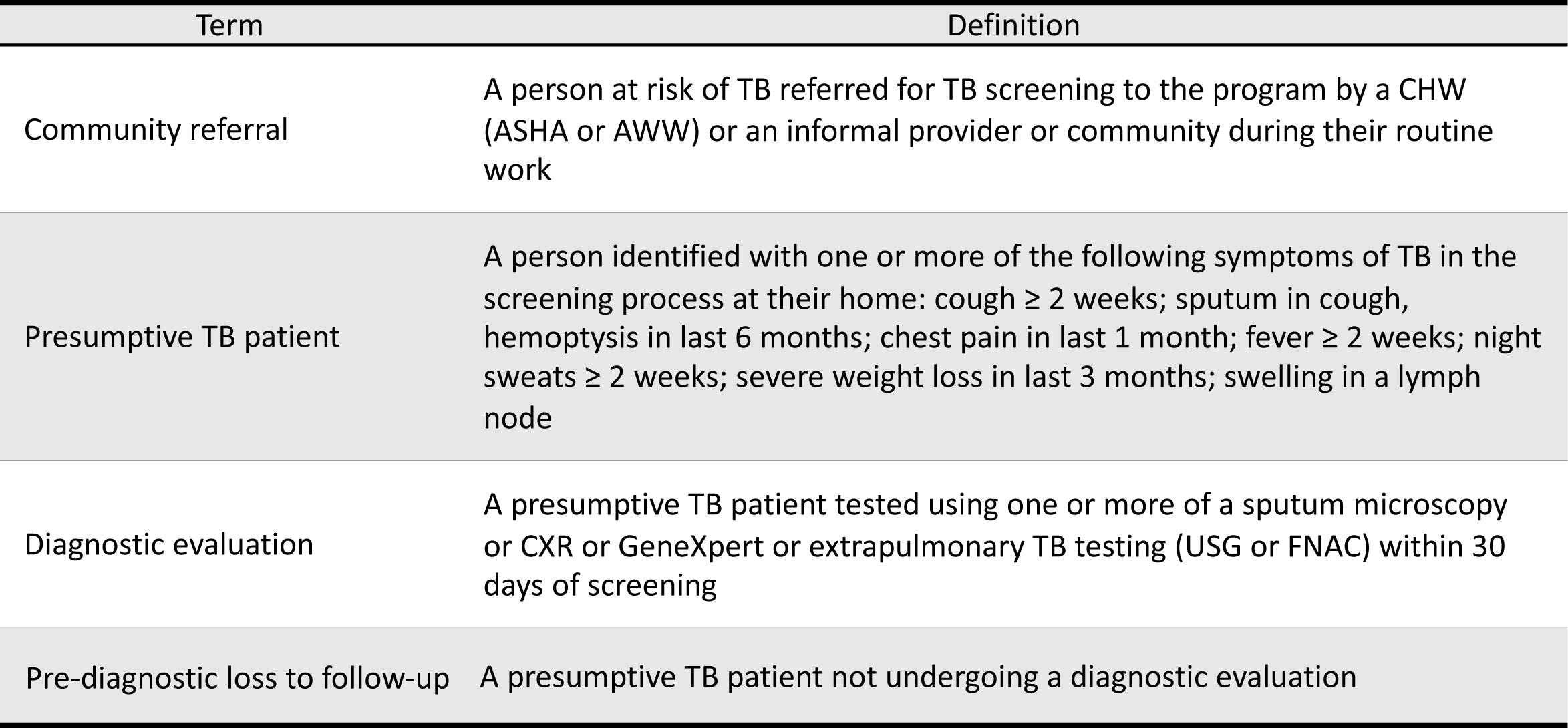
Operational definitions used in the program. CHW: Community Health Worker ASHA: Accredited Social Health Activist AWW: Anganwadi Worker CXR: Chest X-ray USG: Ultrasonography

A conditional incentive of INR 200 (∼USD 3) for community referral and INR 300 (∼ USD 5) for each confirmed diagnosis of TB was given by the project. These incentives to the ASHA were in addition to the RNTCP treatment support incentives. The project’s FCs received a fixed monthly salary.

The project used a Management Information System (MIS) developed in Microsoft Excel which maintained individual patient data electronically. On receipt of a community referral, the FC enlisted them in a referral register with a unique ID and a separate form was completed for screening. The status of each community referral was tracked using the referral register. A patient-wise folder with various forms and diagnostic reports was maintained for all presumptive TB cases undergoing diagnostic evaluation. Data were entered into the MIS weekly and at least 20% entries were verified monthly in a two-person formation to assess data entry quality.

### study population and study period

#### Quantitative

All community referrals in the 3 selected blocks in Samastipur district between January 01, 2018 to December 31, 2018 were included in the study. There was no sampling involved.

#### Qualitative

Presumptive TB cases referred for diagnostic evaluation were interviewed. A total of 7 in-depth interviews were conducted with 3 presumptive TB cases who were evaluated (2 females, 1 male, age range: 17 – 30 years) and 4 presumptive cases who were not evaluated (2 females, 2 males, age range: 6 – 65 years). A purposive sample of ASHAs and field coordinators was selected for the focus group discussions (FGD). Three FGDs were conducted with a group of 8 ASHAs (all females, age range: 24 – 49), 11 ASHAs (all females, age range: 22 – 54), and 9 field coordinators (6 females, 3 males, age range: 20 – 38). The average duration was 36 minutes (range 23 – 51 minutes). A diverse sample of males and females across age groups was purposively selected for diversity and interviewed till saturation was achieved.

### data variables, sources of data and data collection

#### Quantitative

Data on the community referrals’ characteristics (location, referral source, age, gender), screening criteria (date of screening, symptoms, outcome), and diagnostic evaluation (date of test, type of test) were extracted from the MIS into a structured proforma. The data extracted from the MIS was de-identified before being exported for analysis. [Figure 1]

#### Qualitative

Presumptive TB cases were interviewed by TG (a male medical doctor trained in qualitative research) and DS (a female economist trained in qualitative research), and FGDs were conducted by TG and RM (a male professor of social psychology). All the three investigators have long and adequate experience of the socio-cultural context of the region, and understanding of TB programs. After obtaining a participant’s consent, the interviews were conducted at a time and place convenient for the participant and privacy was ensured. The regional vernacular was used for interactions, and audio-recorded using voice recorder after obtaining consent. The objectives of the study were explained to the participants. No participant denied permission for an interview and there were no repeat interviews. A topic guide was used for FGDs and an interview guide for in-depth interviews to explore the enabling factors and barriers for diagnostic evaluation. Appropriate probes were used for clarity and to elicit information. The information was debriefed for participant validation after the interview, but the transcripts were not returned to them.

### analysis and statistics

#### Quantitative

The data were analysed using EpiData Analysis (version 2.2.2.183, EpiData Association, Odense, Denmark) and Stata (version 15.1, StataCorp LLC, College Station, Texas, USA). Patients not undergoing a diagnostic test for TB within 30 days of referral were considered PDLFU. Patients with diagnostic test after 30 days (n = 140) were excluded from the diagnostic evaluation category. In case patients had diagnostic test performed prior to community referral, the time to diagnostic visit was set as 0 days (n = 48). Age was missing for 242 community referrals.[Figure 1] As applicable, variables were summarized with mean (and standard deviation or SD) or median (and inter-quartile range or IQR) based on statistical distribution of data, or frequencies and percentages. Association of demographic and symptom characteristics with diagnostic evaluation was analyzed using χ2-square test and unadjusted relative risk (RR) with 95% confidence interval was calculated. A multivariate analysis using Poisson regression was used to calculate adjusted relative risk (aRR) with 95% confidence interval. P-value ≤ 0.05 was considered statistically significant.

#### Qualitative

The transcripts were prepared on the same day using audio recording and field notes by TG. Manual descriptive content analysis was performed by two independent, trained researchers (TG, DS) to generate categories and themes.[23] Any discrepancies between the two were resolved through discussion. These were discussed and reviewed by RM to avoid subjective bias. The codes and themes were related back to the original data to ensure that the results reflect the data.[24]

### patient and public involvement

Patients were neither involved in the study design nor in the interpretation of patient relevant outcomes. Nonetheless, patient’s views were sought in the qualitative interviews and included in the results. The results of this study will be communicated to the patients and the public through a newsletter in the vernacular.

## RESULTS

### care cascade and characteristics of presumptive TB cases

We received a total of 13395 community referrals, out of which 90.9% (n=12180) were screened for symptoms. Of those screened, 91.5% (n=11146) were presumptive TB cases, and referred for a diagnostic evaluation. (Figure 2)

**Figure 2:**
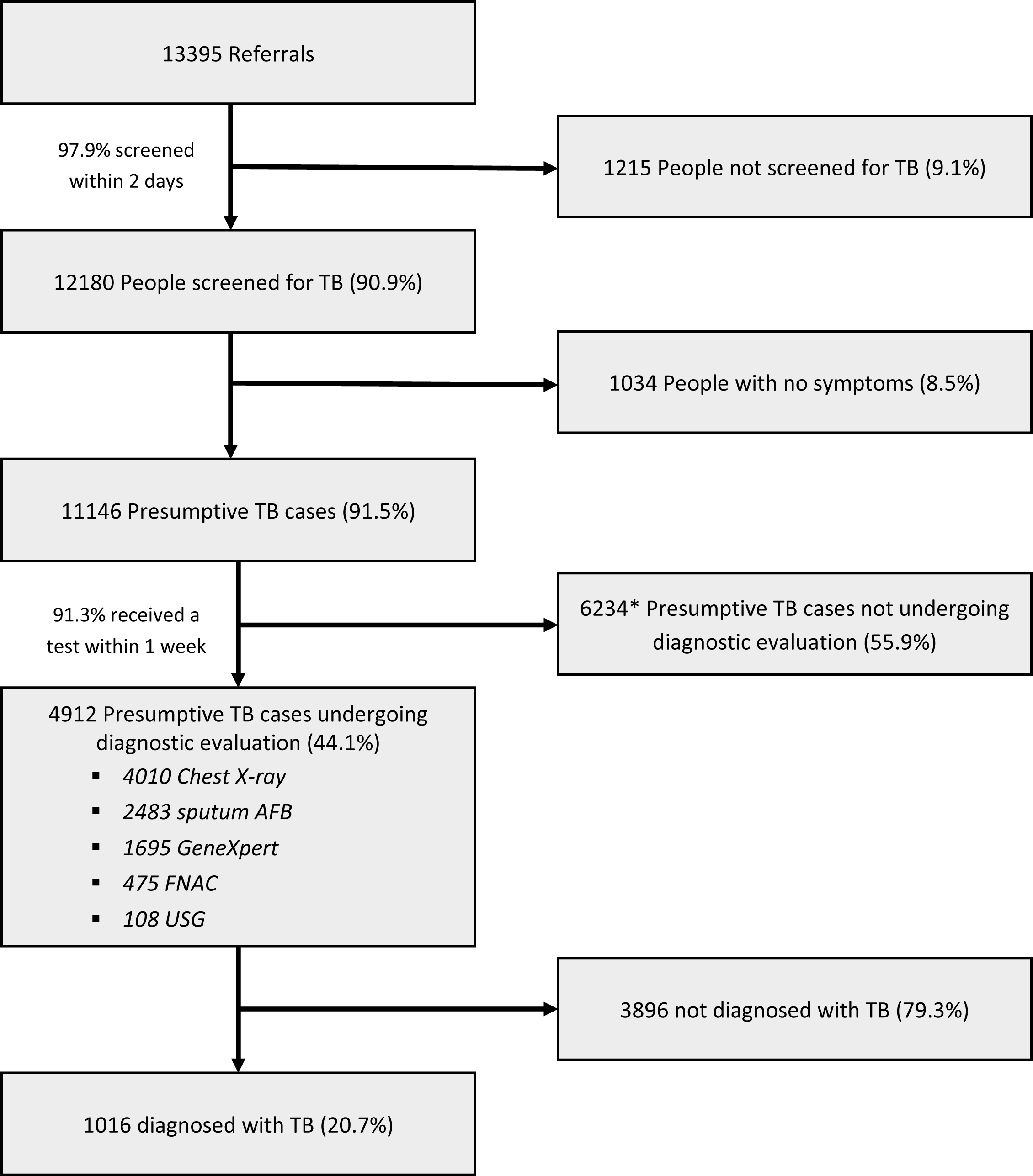
TB care cascade between January 2018 and December 2018 from community referral to diagnostic evaluation in a community-based ACF program in Samastipur, India. All percentages are calculated as a proportion of the number of participants entering the previous step of the cascade. * Includes 140 Presumptive TB cases undergoing diagnostic evaluation beyond 30 days from screening.

There was nearly equal representation of presumptive TB cases from all the 3 blocks, and ASHAs identified most of them (75.6%). The mean age of presumptive TB cases was 35 years with majority in the 15-44 age group (41.8%). There were more males (52.2%) than females. (Table 1)

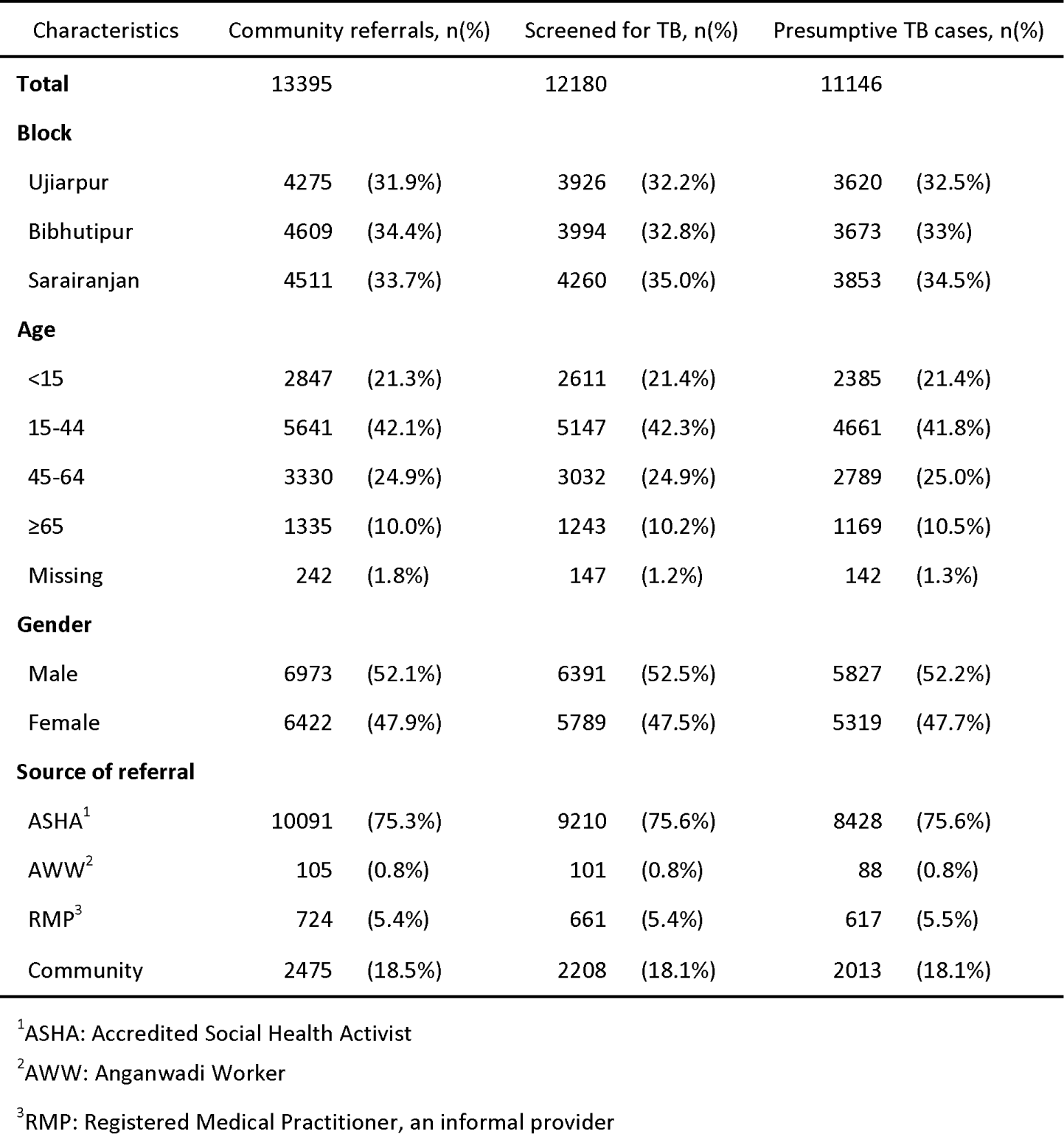

| Characteristics | Community referrals, n(%) |  | Screened for TB, n(%) |  | Presumptive TB cases, n(%) |  |
| --- | --- | --- | --- | --- | --- | --- |
| **Total** | 13395 |  | 12180 |  | 11146 |  |
| **Block** |  |  |  |  |  |  |
| Ujiarpur | 4275 | (31.9%) | 3926 | (32.2%) | 3620 | (32.5%) |
| Bibhutipur | 4609 | (34.4%) | 3994 | (32.8%) | 3673 | (33%) |
| Sarairanjan | 4511 | (33.7%) | 4260 | (35.0%) | 3853 | (34.5%) |
| **Age** |  |  |  |  |  |  |
| <15 | 2847 | (21.3%) | 2611 | (21.4%) | 2385 | (21.4%) |
| 15-44 | 5641 | (42.1%) | 5147 | (42.3%) | 4661 | (41.8%) |
| 45-64 | 3330 | (24.9%) | 3032 | (24.9%) | 2789 | (25.0%) |
| ≥65 | 1335 | (10.0%) | 1243 | (10.2%) | 1169 | (10.5%) |
| Missing | 242 | (1.8%) | 147 | (1.2%) | 142 | (1.3%) |
| **Gender** |  |  |  |  |  |  |
| Male | 6973 | (52.1%) | 6391 | (52.5%) | 5827 | (52.2%) |
| Female | 6422 | (47.9%) | 5789 | (47.5%) | 5319 | (47.7%) |
| **Source of referral** |  |  |  |  |  |  |
| ASHA<sup>1</sup> | 10091 | (75.3%) | 9210 | (75.6%) | 8428 | (75.6%) |
| AWW<sup>2</sup> | 105 | (0.8%) | 101 | (0.8%) | 88 | (0.8%) |
| RMP<sup>3</sup> | 724 | (5.4%) | 661 | (5.4%) | 617 | (5.5%) |
| Community | 2475 | (18.5%) | 2208 | (18.1%) | 2013 | (18.1%) |

<sup>1</sup>ASHA: Accredited Social Health Activist
<sup>2</sup>AWW: Anganwadi Worker
<sup>3</sup>RMP: Registered Medical Practitioner, an informal provider

The most common symptoms among presumptive TB cases were cough ≥ 2 weeks (79.8%%), severe weight loss in 3 months (74.6%), fever ≥ 2 weeks (73.9%), and chest pain in the last 1 month (63.4%), while hemoptysis in the last 6 months (12.1%) was the least common. Nearly one-fifth of the presumptive TB cases reported a previous history of anti-TB treatment (22.4%), and 20.5% were tobacco users. (Table 2)

| Characteristics | Presumptive TB cases, n | Not evaluated, n(%) |  | Unadjusted RR (95% CI) |  | p-value | Adjusted RR (95% CI) |  | p-value |
| --- | --- | --- | --- | --- | --- | --- | --- | --- | --- |
| Total | 11146 | 6234 | (55.9%) |  |  |  |  |  |  |
| Block |  |  |  |  |  |  |  |  |  |
| Ujiarpur | 3620 | 2060 | (56.9%) | 1.0 | (1.0 - 1.0) | 0.87 | 1.1 | (1.0 - 1.1) | 0.00 |
| Bibhutipur | 3673 | 2083 | (56.7%) | Ref |  |  | Ref |  |  |
| Sarairanjan | 3853 | 2091 | (54.3%) | 1.0 | (0.9 - 1.1) | 0.03 | 1.0 | (1.0 - 1.1) | 0.14 |
| **Age** |  |  |  |  |  |  |  |  |  |
| <15 | 2385 | 1625 | (68.1%) | 1.3 | (1.2 - 1.4) | 0.00 | 1.0 | (0.9 - 1.1) | 0.37 |
| 15-44 | 4661 | 2459 | (52.8%) | 1.0 | (1.0 - 1.1) | 0.83 | 0.9 | (0.9 - 1.0) | 0.00 |
| 45-64 | 2789 | 1464 | (52.5%) | Ref |  |  | Ref |  |  |
| ≥65 | 1169 | 588 | (50.3%) | 1.0 | (0.9 - 1.0) | 0.21 | 1.0 | (0.9 - 1.1) | 0.65 |
| **Gender** |  |  |  |  |  |  |  |  |  |
| Male | 5319 | 2859 | (53.8%) | Ref |  |  | Ref |  |  |
| Female | 5827 | 3375 | (57.9%) | 0.9 | (0.9 - 1.0) | 0.00 | 1.0 | (0.9 - 1.0) | 0.00 |
| **Source of referral** |  |  |  |  |  |  |  |  |  |
| ASHA<sup>1</sup> | 8428 | 4690 | (55.7%) | 0.9 | (0.8 - 1.1) | 0.50 | 1.0 | (0.8 - 1.1) | 0.70 |
| AWW<sup>2</sup> | 88 | 52 | (59.1%) | Ref |  |  | Ref |  |  |
| RMP<sup>3</sup> | 617 | 271 | (43.9%) | 0.7 | (0.6 - 0.9) | 0.00 | 0.8 | (0.7 - 1.0) | 0.08 |
| Community | 2013 | 1221 | (60.7%) | 1.0 | (0.9 - 1.2) | 0.77 | 1.0 | (0.9 - 1.2) | 0.91 |
| **Signs and symptoms** |  |  |  |  |  |  |  |  |  |
| Cough ≥ 2 weeks | 8895 | 4514 | (50.8%) | 0.7 | (0.6 - 0.7) | 0.00 | 0.9 | (0.9 - 1.0) | 0.00 |
| Sputum | 6184 | 2779 | (44.9%) | 0.6 | (0.6 - 0.7) | 0.00 | 0.8 | (0.8 - 0.8) | 0.00 |
| Hemoptysis in last 6 months | 1346 | 494 | (36.7%) | 0.6 | (0.6 - 0.7) | 0.00 | 0.8 | (0.8 - 0.9) | 0.00 |
| Chest pain in last 1 month | 7061 | 3260 | (46.2%) | 0.6 | (0.6 - 0.7) | 0.00 | 0.8 | (0.8 - 0.8) | 0.00 |
| Fever ≥ 2 weeks | 8242 | 4289 | (52.0%) | 0.8 | (0.8 - 0.8) | 0.00 | 0.9 | (0.9 - 1.0) | 0.00 |
| Night sweats ≥ 2 weeks | 3730 | 1724 | (46.2%) | 0.8 | (0.7 - 0.8) | 0.00 | 0.9 | (0.8 - 0.9) | 0.00 |
| Severe weight loss in last 3 months | 8318 | 4254 | (51.1%) | 0.7 | (0.7 - 0.8) | 0.00 | 0.9 | (0.9 - 0.9) | 0.00 |
| Swelling in a lymph node | 2067 | 1517 | (73.4%) | 1.4 | (1.4 - 1.5) | 0.00 | 1.0 | (1.0 - 1.0) | 0.96 |
| **Other factors** |  |  |  |  |  |  |  |  |  |
| Previous history of anti-TB treatment | 2501 | 1034 | (41.3%) | 0.7 | (0.7 - 0.7) | 0.00 | 0.8 | (0.8 - 0.9) | 0.00 |
| Alcohol user | 159 | 90 | (56.6%) | 1.0 | (0.9 - 1.2) | 0.86 | 1.2 | (1.1 - 1.4) | 0.01 |
| Tobacco user | 2280 | 1109 | (48.6%) | 0.8 | (0.8 - 0.9) | 0.00 | 1.0 | (0.9 - 1.1) | 0.99 |

<sup>1</sup>ASHA: Accredited Social Health Activist
<sup>2</sup>AWW: Anganwadi Worker
<sup>3</sup>RMP: Registered Medical Practitioner, an informal provider
Age missing for 142 presumptive cases and 98 not evaluated
All % are row percentages.
p-value ≤ 0.05 considered significant

### characteristics of pre-diagnostic LFU and associated risk factors

Nearly 44% (n=4912) of the presumptive TB cases received a diagnostic evaluation. The pre-diagnostic LFU was highest among younger (< 15 year) presumptive TB cases (68.1%), while lesser among those who were referred by an RMP (43.9%) and complained of hemoptysis in the last 6 months (36.7%). On multivariate analysis, presumptive TB cases who were between 15 to 44 years of age (aRR 0.9, 95% CI: 0.9 – 1.0, p = 0) were more likely to be LFU in the pre-diagnostic stage. All of the clinical features of presumptive TB were also associated with lower LFU except lymph node swelling. Alcohol use was an independent risk factor for LFU (aRR 1.2, 95% CI: 1.1 – 1.4, p = 0.01). The median time to diagnosis was 1 day (IQR = 3). (Table 2)

### Enablers to access the first diagnostic evaluation

In the interviews, patients reported the following enablers in getting a diagnostic evaluation: transport allowance for travel to the hospital, free services in the PHS and knowledge of the PHS procedures, accompaniment of ASHA and her assistance in the diagnostic process, and ASHA’s understanding of the PHS functioning. (Figure 3)

**Figure 3:**
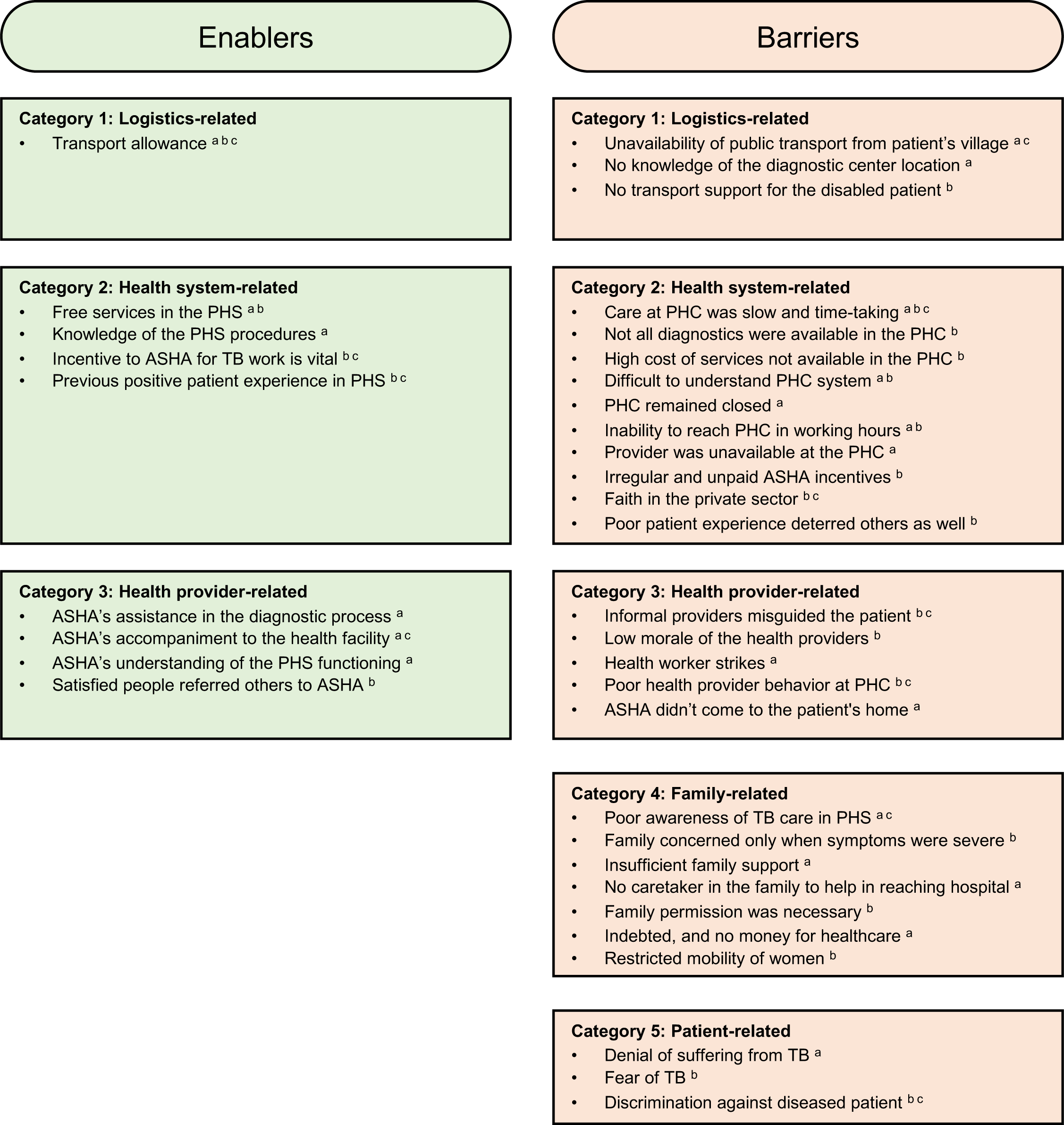
The enablers and barriers in diagnostic evaluation from patients’, ashas’, and field coordinators’ perspective in an ACF TB program in Samastipur, India from January 2018 to December 2018. ^a^: responses of patient ^b^: responses of ASHA ^c^: responses of FC ASHA: Accredited Social Health Activist PHS: Public Health System PHC: Primary Health Centre

*“I started facing financial problems after going to the private hospital*… *Had I known the quality of care in the government hospital earlier, I would have come here. It helped that I didn’t have to spend any money. I went by a vehicle to the PHC and consulted the doctor for free of cost*.*”* (23 years old male patient)

*“I didn’t have to go to the hospital after submitting the sample for diagnostics*… *ASHA took me to the hospital and now sends the medicine*… *I was bony and couldn’t even move a step earlier*… *I became better only from these government drugs; rest were useless. Between RMP and private clinic, we spent nearly ⍰20,000 (∼ USD 292)*.*”* (17 years old female TB patient on medication)

*“When I had chest pain, ASHA told me that we’ll have to go to the PHC, tests will be free, and I’ll get return fare from my home. I thought I’ll get better and decided to go. I got a blood test and x-ray done. Reports were normal and I got some medicines. I felt better*… *If you know the hospital and workers there, it is easier to get things done. Else, you have to spend a lot of time*.*”* (30 years old female patient)

The ASHAs and the Field Coordinators (FCs) expressed transport allowance, incentive to ASHA, positive patient experience, and patient accompaniment as enablers. (Figure 3)

*“If one patient becomes better, she also advises others. Even if we don’t know, the patient who becomes better asks their ill neighbor to call the ASHA for everything. Our mobile number is available in every household*.*”* (ASHA)

*“Sometimes, patients do not have any shared conveyance from their home. In such cases, patients are unable to come, and transport allowance or hiring an autorickshaw for, say 10 patients of the village, really helps. That way, they get consultation and lab tests at PHC, then chest x-ray at another diagnostic facility, and get dropped at their home*.*”* (Field coordinator)

*“ASHA’s income is from incentives, and if she gets incentives in time, she takes up those activities*.*”* (Field coordinator)

### barriers to access the first diagnostic evaluation

Barriers were coded from the interview and FGD transcripts into 5 categories and 27 codes. These barriers are listed in the Figure 3 and described below.

#### Category 1: Logistics-related

As per patients, public transport was not available in certain locations and some patients mentioned their inability to pay for it even where it was available. They also reported not knowing the location of the diagnostic center.

*“I don’t know where the diagnostic center is. From my village, I have to walk for 3 – 4 km before I find any transport*.*”* (65 years old woman)

The ASHA and FC corroborated that transport and insufficient support for certain patient population like disabled people was a key logistics-related challenge.

*“Travel options are also limited for some areas. Either it is not readily available or you have to walk before anything can be found. They don’t have the money to reserve an entire shared vehicle. If the patient is disabled or elderly, they face even more problems*.*”* (Field coordinator)

#### Category 2: Health system-related

According to patients, the care at PHC was slow and took a lot of time. They said either the PHC remained closed or they didn’t reach the PHC in working hours. At other times when they reached, the providers were not available at the facility.

*“I had to go 2-3 times to the PHC before all the tests were completed and reports arrived. Only then I could get the medicines*.*”* (17 years old female patient)

*“We have to run-around a lot in the government hospital. There are huge buildings and it is difficult to figure out what happens where alone*.*”* (30 years old male patient)

The health workers reported unavailability of some diagnostic services in the PHC and the high cost of getting these in the private sector. ASHA’s remuneration for providing TB care services was irregular and unpaid. They indicated that poor patient experience at the PHC also deterred others and that the public put a stronger faith in the private sector.

*“Patient doesn’t know where the registration desk is, where the doctor sits, where the labs and x-ray are done. We have to go with them to get everything completed quickly*.*”* (ASHA)

*“I have provided treatment to 20 patients, but I haven’t been paid for it. It is only in the recent 1-2 years that there has been some improvement. We are not told properly how to fill the document and what to submit for release of payments. All our papers were taken and nothing happened. Earlier we used to only counsel the patient to go for diagnostics, but now we come with them if we get money for helping in diagnosis*.*”* (ASHA)

*“If patient come on a given day and their work doesn’t get done, they won’t come again. Coming again will mean foregoing another day’s wage. They will say that ASHA cheated them and nothing happened at the hospital and nothing is available at the PHC ever. If they go to a private clinic, everything is done in one day and they get the medicine*… *They also prefer private*.*”* (ASHA)

#### Category 3: Health provider-related

Patients described health worker strikes in the field and at the PHC as a barrier. One patient also reported that ASHA doesn’t come to their home.

*“I took the sputum to the PHC, but nothing was done. PHC was on strike for 15 days. The unabated cough and fever didn’t let me sleep. When I felt that I would die, I took an injection from the RMP and drank 5 cough syrups. Only then was there any relief*.*”* (65 years old male patient)

*“What will happen after going to the PHC? I go to the hospital and come back with nothing. Even ASHA doesn’t come. I am going to lose everything in this hustle and bustle*.*”* (35 years old female patient)

The health workers reported RMPs misguiding the patients, low morale of the workers, and poor health provider behavior at the PHC as barriers.

*“RMPs misguide the patients a lot. They give medications and injections without any concern. They are not at risk even if the patient dies. If patient becomes serious and they have made money, they will refer them to hospital. They roam through the villages and people get medicine at their doorstep. Someone who is a laborer, their income for the day is saved*.*”* (ASHA)

*“When we take the patient to the PHC, lab technician will return the sample and asks to come on another day*… *Doctor will also not write any medication and don’t know the updated guidelines… We have to then listen from our patient*.*”* (ASHA)

#### Category 4: Family-related

Patients indicated not knowing the extent of available services in the PHS. Some described severe indebtedness of the family and not having any money for healthcare. According to them, insufficient family support and absence of a caretaker in the family were also barriers.

*“I live with my two granddaughters and my son works as a watchman in New Delhi. The money is irregular. If I’ve no money, I won’t go. In November, I didn’t have money and couldn’t go… For consulting with the RMP, I had to take a loan of ⍰150 (∼ USD 2*.*2). After a week, I’ve been able to return only ⍰50*.*”* (65 years old female patient)

*“Both my kids are alone. My husband has migrated for work. He says that take the kids to a senior doctor in a private hospital. It’ll cost at least ⍰3000 (∼ USD 44) and I am waiting for it… There is only my old mother-in-law in the house besides me. How will I leave them alone at home?”* (Mother of a 6 years old patient)

Health workers mentioned permission from the head of the family to visit facility, restricted mobility of women in the families as compared to men, and family’s concern only if symptoms were severe as barriers.

*“Patients say that we’ll ask the head of the family and come only if they assent. Sometimes they are not sure of the quality of care in the PHS. We also counsel the guardian… This happens more often for women. Men are independent; they don’t have to ask the head or the wife… It is also difficult for women. They have to complete household chores and care for the kids before leaving home for the facility. Often, by the time they reach, the facility will close*.*”* (ASHA)

*“If the symptoms are not severe, then patient takes some medicine from RMP at their home and doesn’t want to come for a test*.*”* (Field coordinator)

#### Category 5: Patient-related

Patients indicated denial of suffering from TB.

*“I don’t need any diagnostics. I didn’t have TB ever in my family. This is all because of gas*.*”* (65 years old patient)

As per health workers, some patients had a fear of TB. They also reported discrimination by the community against TB patients.

*“We don’t say TB at first. If we say it, people hesitate. Some are offended that we have said TB is a possible disease for them. We’ve to tell them gently. One sputum positive patient said that she was hurt that I said she has TB*.*”* (ASHA)

*“A lot of people prefer to keep their disease status hidden. TB is considered contagious and people don’t want neighbors to know of it. Earlier, even patient’s utensils and bedding were kept separate*.*”* (ASHA)

## DISCUSSION

This is the first study to assess the magnitude of the pre-diagnostic LFU in an ACF program along with the barriers and enablers in undergoing a diagnostic evaluation. In our study, nearly 44% of the presumptive TB cases didn’t undergo diagnostic evaluation. Provision of transport allowance for patients to travel for diagnostics, accompaniment and support of ASHA, incentives to ASHA, and word-of-mouth recommendation of satisfied patients emerged as key enablers. The major barriers were challenges in transport despite an allowance, insufficient family and health provider support, inadequate health services, and poor faith in the public sector.

There are strengths of this study. First, we used a mixed-methods design in which the quantitative and qualitative components complemented each other. The qualitative component explored hitherto unresearched reasons for the pre-diagnostic LFU and recorded the perspectives of various stakeholders in the diagnostic process, including patients and CHWs. Second, the study was conducted in an ACF program using routinely collected data and reflects the on-ground reality. Finally, we adhered to the reporting standards: Strengthening the Reporting of OBservational Studies in Epidemiology (STROBE) guidelines for the quantitative part and the COnsolidated criteria for REporting Qualitative research (COREQ) guidelines for qualitative part.[25,26]

There are several limitations as well. First, the study was conducted in a designed ACF program with interventions across the TB care cascade. The site was different than a typical RNTCP implementation and multiple program staff worked on it in addition to the CHWs and RNTCP staff. In addition, the PHS was comparatively weaker in the study region. Second, there was intuitive screening taking place before the patients entered the care cascade. Therefore, the true number of people being screened was not known. Third, we only recorded presumptive TB cases undergoing diagnostic evaluation and there was no recording of presumptive TB cases reaching facility but not undergoing evaluation. Fourth, the yield in terms of confirmed TB diagnosis was known, but we didn’t record the number of people completing the diagnostic process as per the diagnostic algorithm. Fifth, we missed presumptive TB cases accessing TB diagnostic evaluation in the private sector.

The study from the ACF program in Myanmar reported 96.6% of the presumptive TB cases receiving CXR in the mobile van, while only 51.4% of the cases referred for sputum microscopy to the health facility followed through. Immediate availability of a mobile CXR machine was a key factor behind such high rates, and lack of a one-stop diagnostic service was a disincentive for patients.[12] Project Axshya in India reported a low 22% people going to the DMC out of all those referred, while overall 54% received sputum microscopy through sputum transport and referral to DMC combined.[13] In contrast, 41.1% of the presumptive TB cases in our study went to a facility for a diagnostic evaluation. Our qualitative findings corroborate it and suggest that difficulty in transport and poor patient experience are important barriers. A one-stop care center increases the service uptake and patients don’t have to make repeated visits. Similarly, sputum transport mechanisms reduce patient’s hardship. Accompaniment and support of ASHA is a likely reason for this difference with the previous Indian finding.

The Zimbabwe study reported place of residence (rural) and type of facility (private) as important risk factor for pre-diagnostic LFU.[14] While we found association of LFU with various risk factors, their importance in public health programs is limited. However, the qualitative findings point towards important reasons. Previously known challenges in TB programs like TB stigma, patient denial, poor patient experience, and health system deficiencies resonated in our findings. Patients found care seeking in public sector cumbersome, and the informal providers and private sector remained a trusted first choice for most. While there were no significant differences in LFU between the genders, women reported additional challenges of restricted mobility, financial dependence, and burden of household work.

The study has implications for various public health programs targeting improved TB care services, particularly in poor and rural populations. In fact, the barriers in accessing a diagnostic evaluation have a bearing on the access to a broader range of primary healthcare services in similar settings. Learnings from enablers and addressing barriers can improve uptake of the diagnostic services and change the perception of the PHS. We recommend strong patient support mechanism across the TB care pathway, involvement of families in TB care, and systematic reform to address health system-related challenges. Further research in health system challenges involving the perspectives of health system administrators, doctors, nurses, and other health facility workers will inform the process of such a reform.

In conclusion, about three-fifths of the presumptive TB cases were LFU in pre-diagnostic stage and didn’t have a TB diagnostic test. Several enablers and barriers for diagnostic evaluation were identified. If we want to find the missing TB patients, it needs to be easier for presumptive TB patients to navigate the diagnostic process. Health system reforms are rather complex, however, rather simple interventions such as sputum transportation and adequate support from a person knowing the health system may already make big changes.

## Data Availability

The deidentified data used in the study is available from the corresponding author on reasonable request. It contains information on patient demographics and characteristics, screening criteria, diagnostic test used, and final diagnosis. There is no additional data available beyond that.

## ACKNOWLEDGEMENT

We are grateful for the support of RNTCP, Government of Bihar, State TB Officer Dr. (Major) K. N. Sahai, Communicable Disease Officer Dr. Sree Ram Prasad, and District Health Society, Samastipur. We thank Sriram Selvaraju, Anthony D. Harries, Dick Menzies, and participants of the TB Research Methods course at McGill Summer Institute 2018 for their comments on the protocol, and Amol Dongre for his feedback on qualitative results. The program couldn’t have been implemented without the tireless efforts of ASHAs, health providers, and the team at IIH.

This research was conducted through the Structured Operational Research and Training Initiative (SORT IT), a global partnership led by the Special Programme for Research and Training in Tropical Diseases at the World Health Organization (WHO/TDR). The model is based on a course developed jointly by the International Union Against Tuberculosis and Lung Disease (The Union) and Medécins sans Frontières (MSF/Doctors Without Borders). The specific SORT IT programme which resulted in this publication was jointly developed and implemented by: The Union South-East Asia Office, New Delhi, India; the Centre for Operational Research, The Union, Paris, France; Department of Preventive and Social Medicine, Jawaharlal Institute of Postgraduate Medical Education and Research, Puducherry, India; Deartment of Community Medicine and School of Public Health, Postgraduate Institute of Medical Education and Research, Chandigarh, India; Department of Community Medicine, All India Institute of Medical Sciences, Nagpur, India; Dr. Rajendra Prasad Centre for Ophthalmic Sciences, All India Institute of Medical Sciences, New Delhi, India; Department of Community Medicine, Pondicherry Institute of Medical Science, Puducherry, India; Department of Community Medicine, Kalpana Chawla Medical College, Karnal, India; National Centre of Excellence and Advance Research on Anemia Control, All India Institute of Medical Sciences, New Delhi, India; Department of Community Medicine, Sri Manakula Vinayagar Medical College and Hospital, Puducherry, India; Department of Community Medicine, Velammal Medical College Hospital and Research Institute, Madurai, India; Department of Community Medicine, Yenepoya Medical College, Mangalore, India; Karuna Trust, Bangalore, India and National Institute for Research in Tuberculosis, Chennai, India.

## ETHICS APPROVAL

Ethical approval was obtained from Institutional Ethics Committee, Emmanuel Hospital Association, New Delhi (Order 201) and the Ethics Advisory Group, International Union Against Tuberculosis and Lung Disease, Paris (EAG number 98/18). Informed consent was obtained from participants before the in-depth interviews and FGDs.

## AUTHOR CONTRIBUTION

Conceptualization and design: TG, VG, DS, MV, MiB, RM, MaB

Data collection: TG, DS, RM

Data analysis and interpretation: TG, VG, DS, RM

Writing — original draft: TG

Writing — review and edit: TG, VG, DS, MV, MiB, RM, MaB

